# Towards Superhuman Imitation Learning for Sequential Head-and-Neck Cancer Treatment Decisions

**DOI:** 10.64898/2025.12.11.25342119

**Authors:** F. Corna, X. Zhang, G. Canahuate, S. Attia, A. Mohamed, M. Naser, C. Fuller, G.E. Marai

## Abstract

We propose a simulator-driven imitation learning framework for sequential decision making in head and neck cancer (HNC) treatment. Our method, *Superhuman Policy Gradient Optimization (SPGO)*, integrates inverse reinforcement learning principles with policy gradient updates to derive three-stage treatment policies directly from recorded physician decisions. It leverages a pre-trained clinical simulator—combining a variational autoencoder and gradient boosting models—to generate complete, temporally consistent patient trajectories, enabling safe and reproducible training.

Unlike conventional behavior cloning, SPGO optimizes a *sub-dominance loss* that explicitly rewards surpassing the expert across multiple clinical outcomes, including relapse at year three and patient-reported toxicities at multiple follow-up times. We systematically compare six subdominance configurations (absolute vs. relative, sum vs. max aggregation, per-feature vs. max-only *α* updates) to assess how loss design affects convergence and treatment quality.

Our best configuration—relative differences with sum aggregation and per-feature *α* updates—achieves over 70% superhuman dominance across clinically relevant features on held-out patients. The learned policies reproduce expert decisions on acute measures while significantly reducing predicted late toxicities and relapse risk, demonstrating generalization beyond the training distribution.

**CCS Concepts:** • **Applied computing** → **Health informatics**; • **Computing methodologies** → *Reinforcement learning*; *Learning from demonstrations*.

## 1 Introduction

Head and Neck Cancer (HNC) represents a complex clinical domain where therapeutic decisions unfold sequentially across multiple stages, balancing oncological control against long-term toxicity. Although multidisciplinary teams and evidence-based guidelines inform treatment, real-world practice involves heterogeneous patient populations, evolving tumor characteristics, and individualized preferences. This complexity challenges traditional machine learning approaches, which generally predict isolated outcomes rather than optimize entire treatment trajectories.

### Limitations of existing approaches

Purely supervised learning models—such as behavior cloning from physician records—struggle with error compounding: small mistakes at early stages propagate to later decisions, leading to unrealistic or unsafe treatment plans [17]. Reinforcement Learning (RL) can, in principle, handle sequential credit assignment but *requires explicit, scalar rewards*. Unfortunately, healthcare applications usually involve *multiple* clinical objectives (e.g., relapse control vs. toxicity reduction) [15], hence requiring an explicit weighting for combination that is inherently subjective and difficult to align with clinical reasoninge [5].

In contrast, imitation learning does not require such a reward, and simply tries to align the generated trajectory or action with the demonstrations [9]. However, instead of being completely agnostic of the reward, we do have the knowledge of the criteria of interest. So if we could outperform physicians’ demonstrations in terms of all these criteria, we achieve superhuman performance regardless of the combination weights.

Towards this end, we resort to subdominance loss [24] that learns directly from expert trajectories across multiple feature/criterion dimensions, letting the model dynamically reweight each feature through adaptive coefficients [**Ho2016GAIL**, 8]. This allows the policy to discover which objectives are harder to match and to focus optimization there, instead of relying on arbitrary static weights.

### Objective of this work

We leverage *Superhuman Policy Gradient Optimization (SPGO)* [24], an imitation-learning framework that merges inverse RL principles with policy-gradient updates to derive three-stage treatment policies directly from physician demonstrations. Our approach trains on trajectories generated by a high-fidelity simulator [23], enabling systematic experimentation under consistent clinical dynamics. By framing the imitation objective as a *subdominance loss*—which explicitly rewards surpassing the expert across multiple outcomes—we move beyond mimicry towards improved patient trajectories. This formulation does not rely on a manually defined scalar reward but instead derives an adaptive aggregate measure of performance from multiple clinical metrics. The subdominance loss combines feature-wise improvements through either sum or max aggregation, where each feature is dynamically weighted by its adaptive coefficient. In this way, the policy optimizes across multiple outcomes without imposing fixed trade-offs, aligning with the philosophy of focused imitation proposed in recent literature [13]—outperforming demonstrations across features/criteria rather than fitting a static reward.

**Contributions**. We summarize our novelty and contribution as follows:

- Novel integration of a clinical simulator with policy-gradient imitation learning.
- Subdominance loss formulations (absolute vs. relative; sum vs. max aggregation) and adaptive per-feature weighting to focus learning on the hardest clinical outcomes.
- Empirical evidence that SPGO reproduces and often improves on expert decisions, reducing predicted relapse at year three and long-term toxicities.

## 2 Dataset and Simulator

### 2.1 Dataset: MD Anderson Head and Neck Cancer Cohort

Our study uses a de-identified clinical dataset from the MD Anderson Cancer Center (MDACC) comprising 676 patients diagnosed with head and neck cancer (HNC) and treated between 2010 and 2021. Each patient record integrates:

- **Pretreatment features:** demographics (age, sex, smoking), tumor site and stage, HPV status.
- **Treatment decisions: three** sequential treatment stages including definitive surgery (DS), induction chemotherapy (IC), and radiotherapy (RT) with or without concurrent chemotherapy (RT/CC).
- **Patient-reported outcomes (PRO):** 22 symptoms measured longitudinally (Baseline, End of RT, Week 6, Month 3, Month 12) covering pain, fatigue, dry mouth, swallowing, and taste difficulties [16].
- **Treatment Plan Outcomes (TPO):** relapse at year 3, margins, extranodal extension, radiation dose and fractionation [4].

The dataset exhibits missing values and class imbalance, particularly among PRO and long-term relapse data. These limitations motivated the development of a dedicated simulator to generate complete and consistent synthetic trajectories for imitation-learning experiments.

### 2.2 Simulator: VAE + XGBoost Hybrid

To realistically model patient evolution under different *given* treatment policies, we leverage a hybrid simulator that combines generative and predictive components [23].

The simulator reproduces the longitudinal clinical dynamics observed in the MDACC dataset, enabling safe experimentation with learned policies in a controlled, reproducible environment. It models three sequential treatment decisions (*a*_1_, *a*_2_, *a*_3_) corresponding to the standard therapeutic stages in head and neck cancer management. At the first stage, *a*_1_ represents the choice of *Definitive Surgery*; at the second, *a*_2_ corresponds to *Induction Therapy*, where treatment intensity and regimen are determined based on the updated patient state; at the third, *a*_3_ denotes *Radiotherapy or Chemoradiotherapy*, specifying modality, dose, and fractionation parameters. Each decision updates the clinical and symptom variables according to the simulator’s transition function *s*_*t*+1_ = *f*_sim_ (*s*_*t*_, *a*_*t*_), generating a complete trajectory (*s*_1_, *a*_1_, *s*_2_, *a*_2_, *s*_3_, *a*_3_, *s*_4_) that captures both treatment progression and patient outcomes. By decoupling trajectory generation from policy optimization, the simulator provides a stable framework for imitation learning, counterfactual analysis, and longitudinal evaluation of decision policies.

The simulator’s architecture separates (i) *patient generation* at baseline through a variational auto-encoder (VAE [10]) and (ii) *state transition prediction* after each action based on XGBoost [**chen2016xgboost**], enabling controlled, reproducible experiments under a given policy.

1. *Patient Generator (VAE)*. The simulator trained a VAE on pretreatment (baseline) data to learn a latent manifold of heterogeneous patient features, including demographics, tumor descriptors, and baseline PRO. The encoder comprises two fully connected layers (64 units, ReLU activations) followed by mean and variance heads for a 16-dimensional latent space; the decoder mirrors this architecture with separate output heads for numerical (MSE reconstruction) and categorical (cross-entropy) variables. Once trained, the decoder samples *s*_0_ ~(*p*_θ_|*s*_0_)*z* to initialize diverse but clinically plausible baseline states that preserve empirical correlations [14, 11].
2. *Development Predictor (XGBoost)*. For each downstream end-point, a distinct gradient-boosted model [2] is trained that maps (*s*_*t*_, *a*_*t*_) ↦*s*_*t*_+_1_ (or specific components thereof). Categorical targets (e.g., margins, ENE, relapse_yr3) use `binary:logistic or multi:softmax` objectives with macro-F1 or `mlogloss`; numerical targets (e.g., symptom scores) use regression with MSE loss. To address class imbalance, sample weights are computed and a stratified 3-fold cross-validation was used. Training strictly preserves temporal order: for each predicted variable at time *t* + 1, only inputs available up to time *t* are provided (including the current action *a*_*t*_), thereby avoiding target leakage and preserving clinical causality.

#### Symptom Burden Model (SBM)

Patient-reported outcomes (22 MDASI-HN symptoms) are collected at Baseline, End of RT, Week 6, Month 3, and Month 12 [16]. To obtain a compact, robust representation and to address missing values, a Symptom Burden Model (SBM) is employed that labels each timepoint as *high* or *low* burden according to validated severity rules: (i) any symptom ≥7 at that timepoint, or (ii) at least two symptoms ≥5 [18, 12]. These binary clusters, optionally complemented by imputed PRO trajectories in a latent symptom space, reduce dimensionality and noise while maintaining longitudinal consistency. The resulting burden features feed both the VAE (conditioning baseline generation) and the XGBoost-based transitions.

### 2.3 Dataset Used in This Work

#### Data preprocessing

Missing data arise naturally in longitudinal medical records due to incomplete follow-up, unrecorded measures, or variations in clinical documentation. To maintain temporal and structural consistency across datasets, missing values were imputed using a k-Nearest Neighbors (KNN) imputer (k = 3 or 5) for the MDACC-ALL dataset, ensuring completeness before model training. For the MDACC-SBM dataset, imputation was handled internally by the SBM API, producing fully populated symptom trajectories. In the clinical simulator, missing information in intermediate states is automatically re- constructed through internal predictive models based on Variational Autoencoders (VAE) and XG-Boost regressors, ensuring that each simulated trajectory remains temporally coherent and biologically plausible.

#### Dataset use and split

We employ a curated variant of the MDACC– SBM dataset, obtained after filtering out variables with inconsistent values or unreliable statistical distributions. The complete dataset includes:

- 16 pretreatment variables (*Age, Gender, Ethnicity, Race, Smoking Status, Tobacco History, Tumor Site, AJCC Stage, Laterality, T/N/M Stage, HPV Status, Enrollment Status*);
- 3 treatment action variables (*Definitive Surgery, Induction Chemotherapy, Radiotherapy/Chemoradiotherapy*);
- 5 post–intervention features (*ENE, Margin, RT Type, RT Dose, RT Fraction*);
- 20 PRO features (4 symptoms × 5 timepoints);
- 1 long–term treatment outcome (*Relapse at Year 3*).

The resulting dataset is compact yet clinically informative, well suited for imitation learning, reinforcement learning, and counterfactual policy evaluation. Finally, the dataset is split into **80% for training** and **20% for testing** to ensure reliable generalization and performance assessment.

#### Advantages for imitation learning

Due to ethical reasons, it is impractical to directly apply the learned policy to real patients. This challenge plagues many policy evaluation tasks in an offline setting [21]. The simulator provides a convenient and efficient solution which:

- Ensures complete trajectories (no missing values) and stable dynamics for policy training.
- Enables evaluation of multiple subdominance loss configurations under identical simulated conditions.
- Provides a safe offline environment for exploring treatment policies beyond physician demonstrations.

## 3 Methodology

### 3.1 Overview

We model the head and neck cancer treatment pathway as a three-stage sequential decision process, illustrated in Figure 1. At each stage *t* ∈ {1, 2, 3}, the environment provides the current patient state *s*_*t*_, representing demographic, clinical, and symptom features. The corresponding policy network 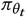 produces a treatment decision 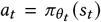, which is executed within the simulator. The simulator then updates the patient state to *s*_*t*+1_ = *f*_sim_ (*s*_*t*_, *a*_*t*_), capturing the clinical evolution under that action. Implicitly inside *f*_sim_ we carry over *s*_*t*_ and *a*_*t*_. This process repeats across the three stages, producing a full trajectory *ξ* = (*s*_1_, *a*_1_, *s*_2_, *a*_2_, *s*_3_, *a*_3_, *s*_4_) [**Ng2000IRL, Ho2016GAIL, Sutton2018RLBook**, 7].

**Figure 1:**
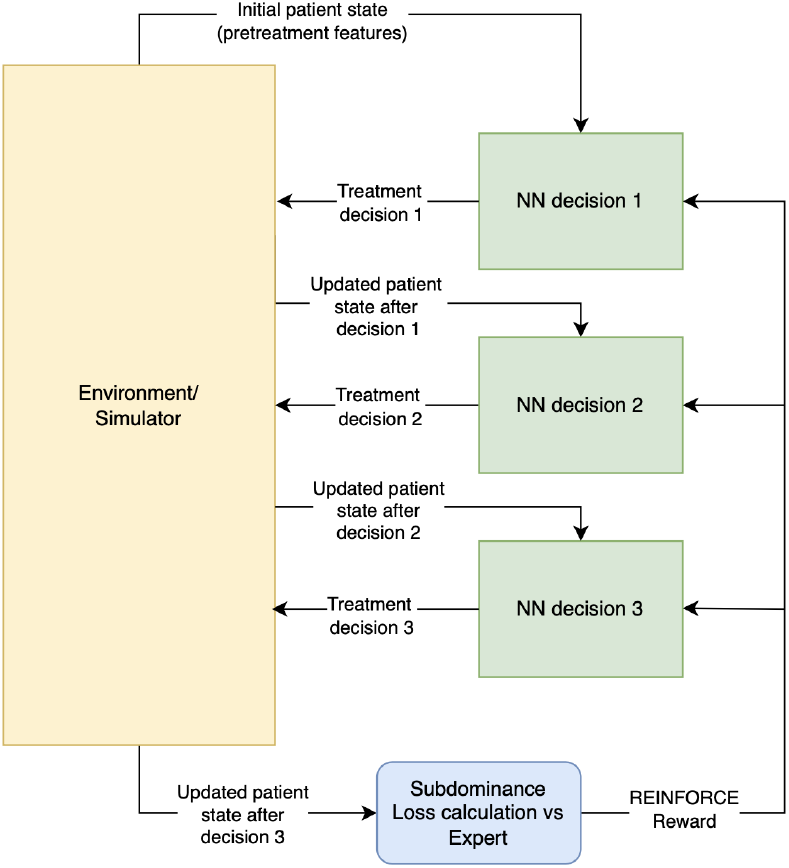
Overview of the architecture that handle the SPGO training cycle.

**Figure 2:**
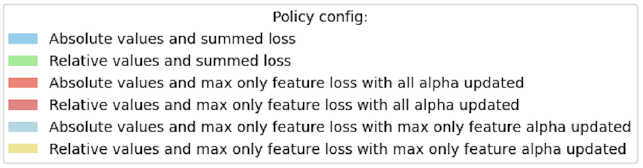
Comparison of six subdominance configurations.

Our proposed framework, *Superhuman Policy Gradient Optimization (SPGO)* [24], integrates inverse reinforcement learning principles with policy-gradient updates [**Finn2016GuidedCost**, 22]. Unlike standard reinforcement learning, which relies on a scalar reward, SPGO derives a multi-objective reward from a *subdominance loss* that quantifies feature-wise improvement over expert trajectories. This enables learning directly from demonstration data without requiring explicit reward design [3, 20].

### 3.2 Three Policy Networks

We train three distinct policy networks corresponding to the sequential medical decisions across the treatment pathway. Each policy network operates conditionally on all prior information available at its decision stage, thus capturing the causal and temporal dependencies of the clinical process.

- **Action1PolicyNN:** receives baseline patient features, including demographics, tumor characteristics, and baseline symptom burden (from the SBM). It outputs a binary decision (e.g., definitive surgery vs. induction therapy).
- **Action2PolicyNN:** takes as input all features from the previous stage, the decision made by *Action1PolicyNN*, and the updated patient state produced by the simulator after executing that decision. This concatenated representation reflects both historical and new clinical information.
- **Action3PolicyNN:** receives the full trajectory history up to stage 2—including all previous states, decisions, and simulated updates—and outputs the final treatment choice among three alternatives (e.g., RT, RT+CC, or observation).

Formally, each policy 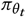 maps *s*_*t*_ to a probability distribution over actions:

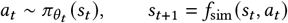

here *f*_sim_ denotes the simulator that generates the new clinical features conditioned on the chosen treatment. Thus, the full simulated trajectory *ξ* = (*s*_1_, *a*_1_, *s*_2_, *a*_2_, *s*_3_, *a*_3_, *s*_4_) encodes the entire decision-making process and its resulting patient features [**Ng2000IRL, Ho2016GAIL, Finn2016GuidedCost, Sutton2018RLBook**].

### 3.3 Subdominance Loss

SPGO compares the agent trajectory *ξ* and the expert trajectory 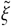 across *K* criteria computed at the final stage, with each *f*_k_ (*ξ*) (and 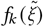) implicitly depending on the full sequence of states and actions that led to those outcomes (e.g., PROs at ENDRT/WK6/M3/M12 and `relapse_yr3`). Letting *f*_*k*_ (*ξ*) be the cost or measure under criterion *k*, we will refer to it as **feature** *k* by following the terminology in [24]. Then the *subdominance* loss is defined as

#### Absolute form

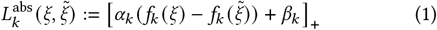

#### Relative form

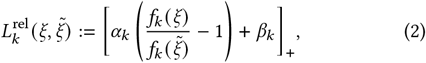

where [*x*]_+_ = max {*x*, 0, *β*_*k*_} is a target margin, and *α*_*k*_ is an adaptive slope updated online. These per-feature losses can be aggregated through one of the following formulae:

#### Summation to promote global improvement

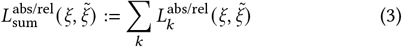

#### Max to focus on the worst violation

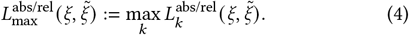

Given a set of demonstration trajectories 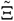, i.e., existing patient’s treatment recording, we define the stochastic reward as

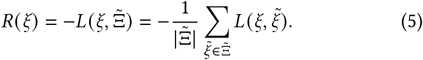

Here we simplified the notation on *L*, which can have four instantiations from abs/rel × sum/max. We will include sub/superscripts such as *k, α, β*, abs/rel, sum/max, whenever the context needs more clarity.

### 3.4 Training Strategy

We trained the policy by using the REINFORCE update [22], where the reward is served by the negative of subdominance loss. Training proceeds online at the patient level with batches of 10 patients. Gradients are accumulated over the batch of *π*_*θ*_-generated trajectories ℬ and backpropagated jointly through all policy networks:

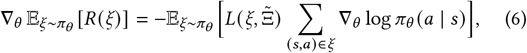

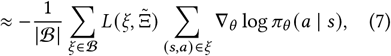

where each *ξ* ∈ ℬ is drawn from *π*_θ_. Noting that 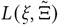 is defined by averaging over the entire demonstration set from Eq (5), we can further approximate it by using just the single trajectory that corresponds to the same patient as *ξ*. That means they share the same initial state *s*_1_.

When sampling the patients to form the batches ℬ, we ensure that, within each iteration, each patient appears in one and only one batch. Gradient clipping (max-norm = 1.0) prevents exploding gradients, and the Adam optimizer (10^−3^ learning rate) ensures stable convergence. The whole procedure is summarized in Algorithm 1.

Four configurations are available for testing:

- **Absolute** vs. **Relative** difference, i.e., Eq (1) vs. Eq (2);
- **Sum** vs. **Max aggregation**, i.e., Eq (3) vs. Eq (4);

By default, all *α*_*k*_ parameters are jointly updated at each training iteration, allowing the model to continuously adjust its sensitivity to each outcome feature. This mechanism dynamically redistributes learning focus according to the observed deviations in subdominance. However, when using the *max* aggregation strategy, simultaneous updates of all *α*_*k*_ can lead to excessive reactivity to minor fluctuations. To mitigate this, we instead test a selective variant in which only the parameter 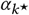 associated with the largest subdominance term is updated at each step. Such a **Max-only** *α*_*k*_ adaptation is applied under the *Max aggregation* configuration, highlighting how concentrated updates can stabilize convergence when optimization is dominated by a single critical feature. *Sum aggregation*, instead, is compatible with both **Update-all** and **Max-only** *α*_*k*_ adaptation. So in total, we tested **six** configurations.

### 3.5 Stability and Interpretability

We applied the following techniques to improve the training stability and the interpretability of the learned model:

- Logit clipping and temperature-based sampling stabilize exploration during early training.
- Adaptive *α*_*k*_ coefficients redistribute gradient pressure toward unsatisfied objectives, forming a dynamic curriculum.
- Integration with the Symptom Burden Model provides low-noise, complete trajectories that enhance stability.
- Monitored metrics include mean reward, *α*_*k*_ evolution, and superhuman dominance rate.

## 4 Experiments on Superhuman Imitation

Evaluating a policy is difficult because it is unethical to directly apply a learned policy to real patients, a typical challenge in offline policy evaluation [21]. We therefore used the simulator for evaluation. Even though the test set does carry the outcome from physician’s decision, we still used the simulator to roll out the progression, hence factoring out the possible mismatch between the simulator and reality, ensuring a fair comparison between the learned policy and physician’s decision.

### Algorithm 1 Superhuman Policy Gradient Optimization (SPGO) Training Loop

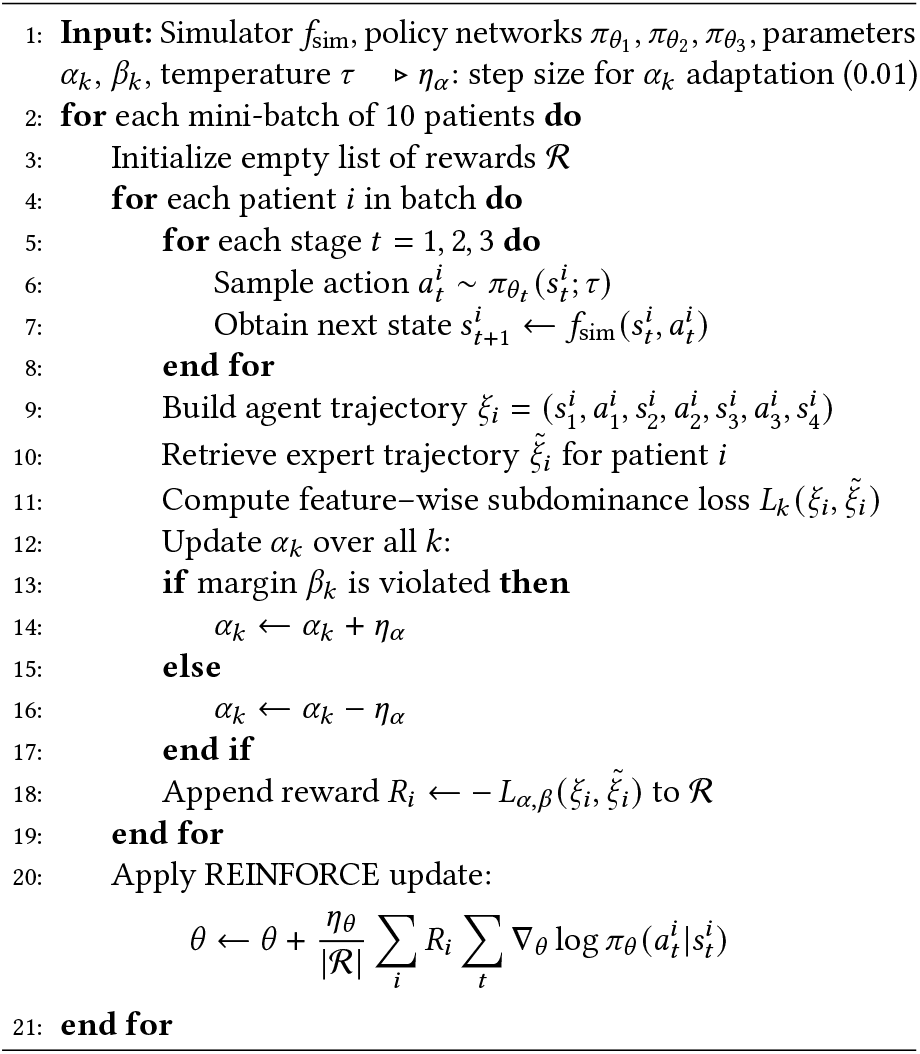

### 4.1 Overall dominance

We tested six configurations of the subdominance loss (absolute vs. relative differences; sum vs. max aggregation; update-all *α* vs. max-only *α*) as defined in Section 3.

Because Superhuman Policy Gradient Optimization (SPGO) is designed to surpass expert decisions rather than merely reproduce them, traditional metrics that compare the predicted action with the doctor’s real decision are insufficient. We then focus on how our policies “dominate” the physician’s policy, evaluating both the sum of all final features 3 and the number of dominated features 4.

#### Dominance across aggregated toxicity Fig. 3

For each patient, the total toxicity is computed as the sum of all Patient-Reported Outcomes (PROs) and Treatment Plan Outcomes (TPOs) obtained from the simulator rollouts. The policy achieving the lower total toxicity value is considered dominant. Across various configurations of the subdominance loss, the SPGO policy achieves dominance in the majority of cases (approximately 61–74%), with the highest rate (73.72%) obtained using the *relative summed loss* formulation. This suggests that optimizing for relative improvements over the expert baseline yields better generalization and overall symptom reduction. Configurations based on absolute or max-only losses exhibit slightly lower dominance, indicating a trade-off between stability and optimization strength.

**Figure 3:**
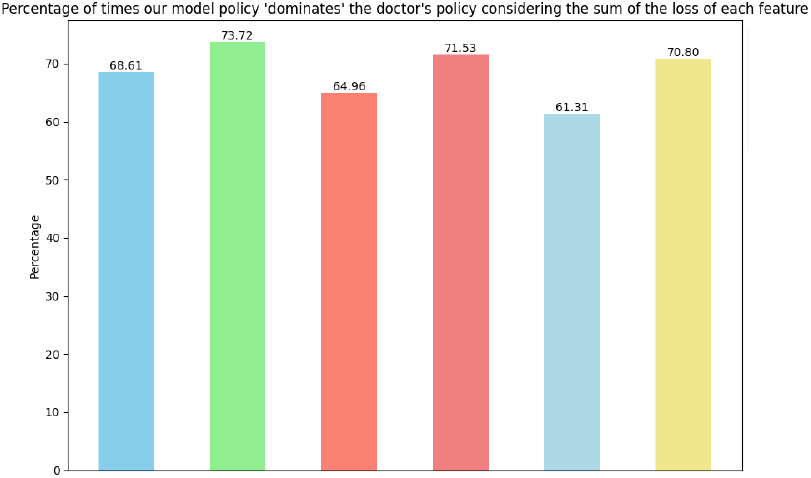
Percentage of patients where our policy dominates the expert when summing feature-wise subdominance losses.

#### Dominance by majority of features Fig 4

In a complementary analysis, dominance is computed feature-by-feature rather than from total toxicity. For each patient, SPGO and physician outcomes are compared across all PRO and TPO features; the policy that yields lower values in more than half of the features is deemed dominant. This majority-based metric confirms SPGO’s superiority, with dominance observed in roughly 70% of patients under relative-loss configurations and above 60% under absolute ones. The consistency across both dominance metrics demonstrates that the learned policy systematically improves on the expert’s behavior, maintaining robustness and generalization across different clinical aspects.

**Figure 4:**
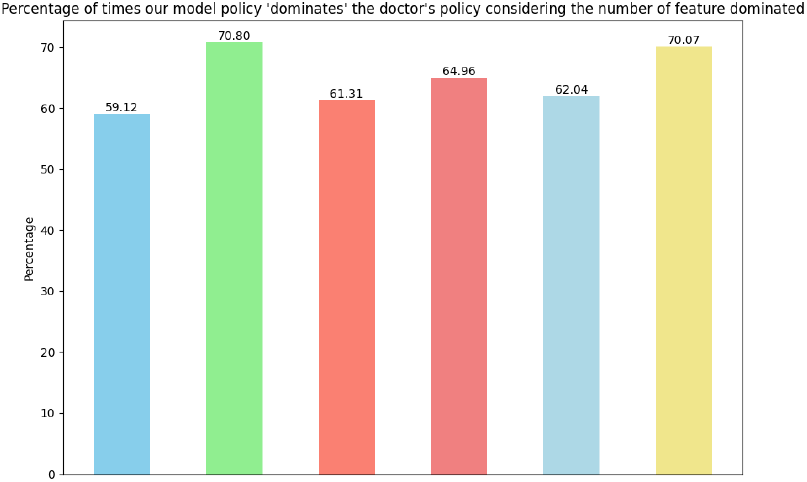
Dominance rate computed by majority of features across patients.

**Performance:** 73.7% dominance across all features jointly and ≈ 70.8% dominance by majority of features on the held-out test set.

**Best configuration:** relative differences + sum aggregation + per-feature *α* updates. As such, we will henceforth only report the results obtained from this configuration.

### 4.2 Training Dynamics

#### Reward Curve

Figure 5 illustrates the evolution of the batch–averaged reward, defined as the negative mean of the subdominance loss multiplied by the sum of log–probabilities, across training iterations. The reward represents the inverse of the batch–level subdominance loss, with an optimal value of 0 corresponding to trajectories that fully dominate the expert. The dashed green line marks the nominal dominance margin *β* = 1, which in the subdominance formulation defines the minimum improvement required for the policy to surpass the demonstrator. However, in the relative loss formulation, the true dominance threshold depends on both *α*_*k*_ and *β*_*k*_, approximately following:

**Figure 5:**
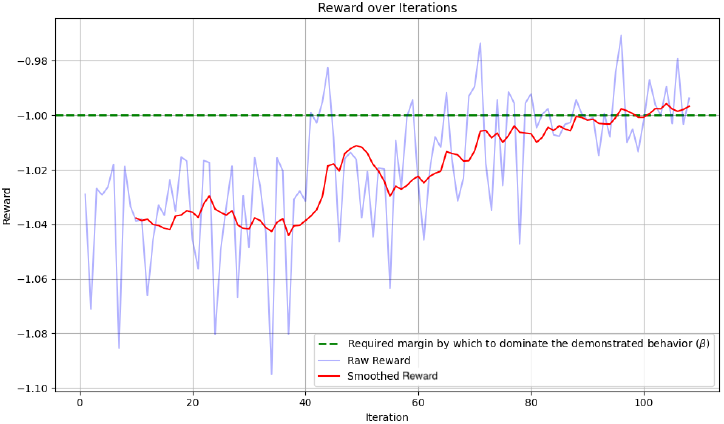
Reward curve showing convergence beyond dominance margin *β*.

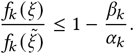

Since *α*_*k*_ values are updated online for each patient rather than per iteration, and vary feature–wise, this fixed line serves only as a qualitative reference. As training progresses, the smoothed curve rises steadily toward 0, indicating a progressive reduction of sub-dominance violations and a consistent improvement of the learned trajectories over the expert baseline. This trend is consistent with the gradual curriculum-learning behavior expected from adaptive *α*_*k*_ weighting [1, 6]

#### Evolution of *α*_*k*_

The adaptive per-feature coefficients exhibit a monotonic decay from their initial value of 1.0 (Fig. 6). Acute features (End of RT, Week 6) decrease slowly, while Month-3/12 and symptom groups show an intermediate decay. The *relapse* coefficient decays the fastest and ends at the lowest level among all features, indicating that its margin is satisfied early; after that point, the curriculum shifts gradient pressure toward the remaining outcomes whose margins are still active.

**Figure 6:**
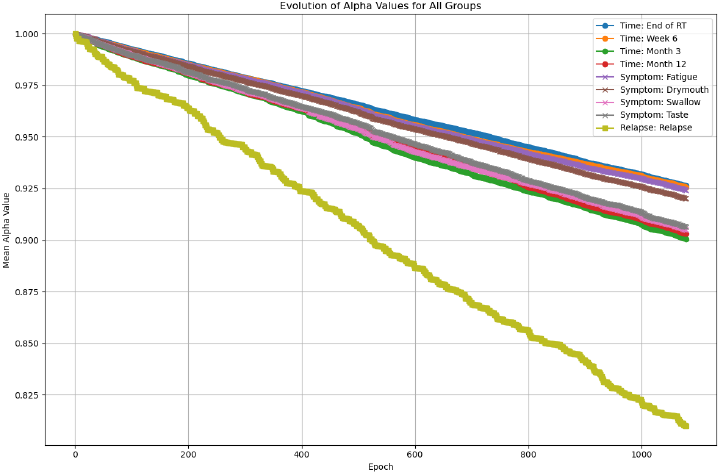
Evolution of *α*_*k*_ coefficients during training, showing adaptive reweighting.

### 4.3 Clinical Outcomes on Test Patients

We trained the SPGO policy independently across seven different dataset partitions, each obtained through random reshuffling of the patient set. This procedure ensures exposure to varied patient combinations and enables assessment of the model’s robustness and stability under different sampling conditions.

#### Relapse at Year 3

The policy exhibits consistent behavior across different random train–test splits, stably leding to an average of 54.04% non-relapse cases with a variance of 2.5%. This robustness to data partitioning confirms the stability of the learned policy and its ability to generalize beyond the specific training configuration, while preserving clinical interpretability [19].

#### Toxicity Over Time

Our policy yields to toxicity values that declines from End of RT to Month 12, mirroring realistic recovery patterns observed in clinical follow-up data.

#### Toxicity by Symptom Domain

Under our policy, higher toxicity values are observed in the *Dry Mouth* and *Taste* domains compared to the *Fatigue* and *Swallowing* domains.

### 4.4 Comparison with Physician

SPGO policies were first evaluated against the expert trajectories contained in the dataset. Figure 7 shows outcomes grouped by time-point (End of RT, Week 6, Month 3, Month 12), showing nearly overlapping toxicity at treatment end but lower values for SPGO in later stages, indicating improved long–term control without loss of acute effectiveness. Figure8 shows outcomes grouped by symptom type (Fatigue, Drymouth, Swallow, Taste), where SPGO closely tracks the physician overall but shows clear reductions in swallowing and taste toxicities, reflecting its finer adjustment to late–onset side effects. This trend supports the central hypothesis that SPGO reaches beyond pure imitation by improving upon clinically critical endpoints [3, 20], demonstrating its dominance across toxicity-related features.

**Figure 7:**
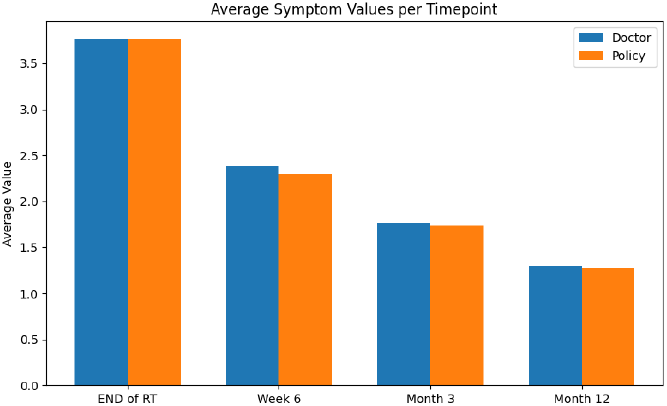
Comparing SPGO with physician’s policy in terms of average toxicity values averaged over multiple symptom domains, grouped by timepoints.

**Figure 8:**
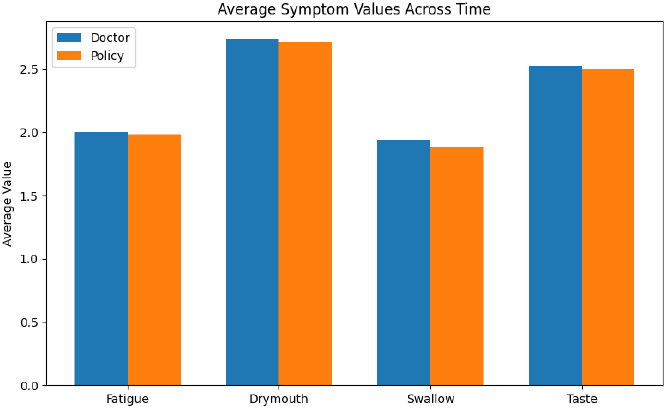
Comparing SPGO with physician’s policy in terms of average toxicity values averaged over multiple follow-up timepoints, grouped by symptom domains.

#### Relapse agreement and net improvement

Figure 9 shows the joint distribution (in %) of relapse outcomes across the two policies. The model reproduces the physician’s relapse pattern with an overall agreement of **85.4%**, obtaining same outcome features for non-relapse and relapse cases in 51.09% and 34.31% of patients, respectively. The off-diagonal entries quantify the disagreement: in 5.84% of test patients, the physician’s policy yielded no relapse while SPGO yielded relapse (*underperforming*); conversely, in 8.76% of cases, the SPGO policy yielded no relapse while the physician’s did not (*outperforming*). Since 8.76% *>* 5.84%, more patients are shifted from relapse to non-relapse by SPGO than the reverse, yielding a **net improvement of** + 2.92 **percentage** of patients in simulated relapse control. This indicates that SPGO maintains clinical plausibility while surpassing the physician on the relapse dimension under the simulated dynamics.

**Figure 9:**
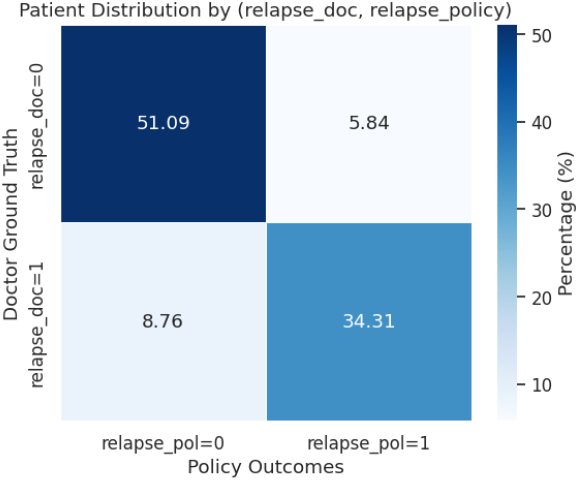
Joint distribution of relapse outcomes for physician and SPGO policies. SPGO maintains overall agreement (85.4%) while achieving a net +2.92% gain in non-relapse outcomes.

### 4.5 Comparison with Behavioral Cloning

To further compare with imitation-based learning, we trained a standard *Behavioral Cloning* (BC) model [**Ho2016GAIL**]. Each BC network mirrors the architecture of the corresponding SPGO policy (three-layer MLP with ReLU activations and dropout) but is trained in a purely supervised manner using cross-entropy loss between predicted and expert actions. Input states include all features available at each decision stage (pretreatment data, previous decisions, and simulated symptom-burden states). Training proceeds with early stopping on validation accuracy and without any interaction with the simulator.

When evaluated under the same simulated environment, the BC policy reproduces the physician’s sequence of actions with high fidelity and led to similar final stage features but fails to improve long-term clinical outcomes.

#### Toxicity comparison with BC

Figure 10 shows that SPGO consistently achieves lower simulated toxicity values compared to those obtained under the BC policy, confirming that the policy-gradient mechanism guided by the subdominance loss enables targeted improvement rather than mere replication of expert behavior (as BC does).

**Figure 10:**
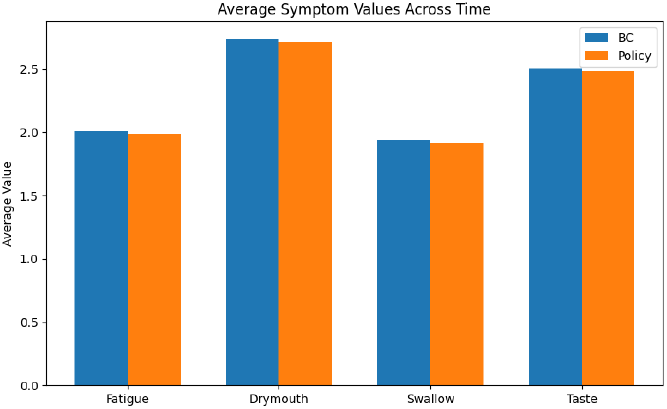
SPGO vs BCpolicy average toxicity values over multiple follow-up timepoints, grouped by symptom domains.

#### Relapse comparison with BC

We further compared the relapse predictions between SPGO and the BC baseline.

Figure 11 reports the joint distribution (in %) of relapse outcomes obtained under BC and SPGO. The two models produced the same outcome in 83.95% of patients (46.72% no-relapse, 37.23% relapse). The off-diagonal entries quantify the disagreement: in 6.57% patients, the BC policy led to no relapse while SPGO led to relapse (*underperforming*); conversely, in 9.49% of cases, the SPGO policy led to no relapse while the BC did not (*outperforming*). This indicates that SPGO also surpasses BC on the relapse dimension under the simulator dynamics.

**Figure 11:**
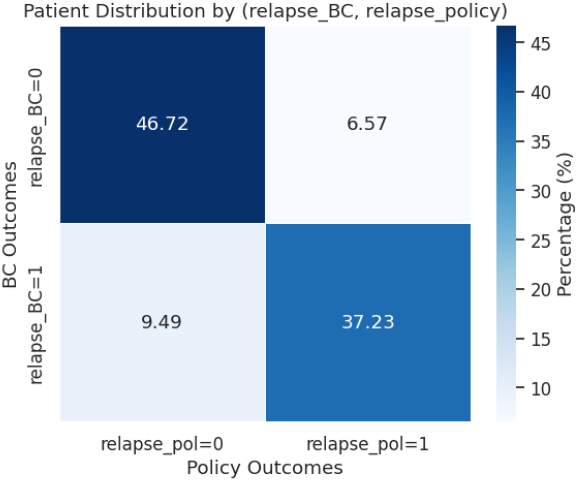
Joint distribution of relapse outcomes for BC and SPGO policies. SPGO maintains overall agreement (83.95%) while achieving a net +2.92% gain in non-relapse outcomes.

Figure 12 compares BC and SPGO through per–toxicity differences Δ = BC −SPGO. Distributions are mostly centered around zero with a slight positive bias, indicating that SPGO achieves lower (thus better) toxicity values, particularly at later timepoints and for swallowing–related metrics. These results suggest that SPGO consistently yields improved long–term outcomes while preserving variability, confirming that its advantage arises from effective optimization rather than stochastic effects.

**Figure 12:**
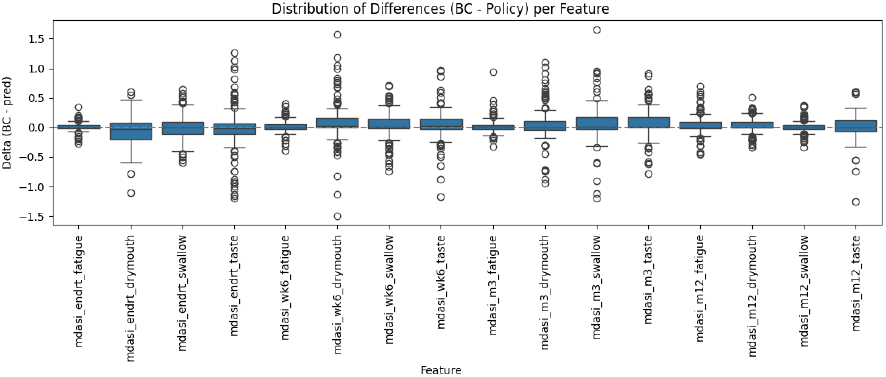
Boxplots shows the distribution of differences between BC and Learned SPGO Policy outcomes. Positive values indicate the SPGO policy achieves lower (better) outcomes.

## 5 Conclusion and Future Work

This work introduced *Superhuman Policy Gradient Optimization (SPGO)*, an imitation-learning framework for sequential cancer treatment planning that integrates policy-gradient reinforcement learning with a clinically grounded subdominance loss. Trained on the MD Anderson Head and Neck Cancer dataset, SPGO learns three-stage treatment policies that replicate expert behavior in early decisions while surpassing physicians and supervised baselines in long-term outcomes. By optimizing feature-wise dominance rather than a handcrafted scalar reward, SPGO aligns model learning with multidimensional clinical objectives—reducing predicted late toxicities and relapse risk.

Experimental results demonstrate consistent improvement across multiple outcome domains. SPGO achieves lower simulated toxicity outcomes, maintains realistic acute decisions, and exhibits a net positive shift in relapse prediction relative to both physicians and behavioral cloning baselines. These findings confirm the potential of subdominance-based policy gradients to bridge the gap between explainable imitation learning and outcome-oriented reinforcement learning in healthcare decision-making.

### Future work

Several research directions follow naturally from this study:

- **Expanded simulation fidelity:** extending the VAE–XGBoost simulator to model competing risks, time-to-event dynamics, and treatment side effects jointly, thereby improving causal realism.
- **Continuous-action and uncertainty-aware policies:** replacing discrete treatment classifiers with Gaussian policies or Bayesian ensembles to represent therapeutic intensity and confidence.
- **Human-in-the-loop refinement:** integrating clinician feed-back during policy updates to ensure safety and maintain interpretability in real-world deployment.
- **Cross-domain validation:** applying SPGO to other sequential clinical decision problems (e.g., radiotherapy fractionation, multimodal oncology, or chronic disease management) to assess generalizability.

In summary, SPGO demonstrates that imitation learning enriched by dominance-aware policy gradients can move beyond replication toward measurable clinical improvement, providing a scalable foundation for the next generation of data-driven decision-support systems in personalized medicine.

## Data Availability

All data produced in the present study are available upon reasonable request to the authors

## Acknowledgment

This research is supported by NSF Award #2312955 and NIH Award #1 R01 CA258827-01.

